# The SMART-AI trial: Real-time cholangioscopy artificial intelligence for the classification of biliary strictures

**DOI:** 10.64898/2025.12.17.25342271

**Authors:** Neil B. Marya, Patrick D. Powers, Matthew Marcello, Prashanth Rau, Navine Nasser-Ghodsi, Christopher Marshall, Jaroslav Zivny, Jad P. AbiMansour, Vinay Chandrasekhara

## Abstract

**Background:** Sampling techniques have poor accuracy for classifying biliary strictures as benign or malignant. Previously, a cholangioscopy artificial intelligence (AI) outperfromed sampling techniques based solely on analysis of previously recorded cholangioscopy footage. The aim of this trial was to compare the performance of a real-time cholangioscopy AI to both sampling techniques and human observers for the task of biliary stricture classification.

**Methods:** A cholangioscopy AI computer connected directly to a cholangioscope console. The computer analyzed the cholangioscopy video stream during procedures for suspected biliary strictures.

The primary outcome of the study was comparison of the performance of cholangioscopy AI to sampling techniques – brush cytology and transpapillary forceps biopsy – for biliary stricture classification. Secondary outcomes included comparison of the AI classification performance to that of human observers (separated into junior-level and experienced-level cohorts) who reviewed the cholangioscopy footage.

**Results:** A total of 41 patients were enrolled in the trial and had biliary strictures analyzed by cholangioscopy AI. For the classification of strictures, the AI had greater classification accuracy than standard sampling techniques (87.8% versus 67.4%; p = 0.043). Additionally, the cholangioscopy AI was significantly more accurate for biliary stricture classification than both junior-level (87.8% versus 61.5%; p = 0.001) and experienced endoscopists (87.8% versus 63.15%; p = 0.011).

**Conclusions:** This trial demonstrates that sampling techniques and human assessment of biliary strictures are flawed and there may be a benefit to the use of a cholangioscopy AI system to aid in biliary stricture classification.

## BACKGROUND

Cholangiocarcinoma (CCA) is an aggressive tumor that results in biliary strictures and subsequent biliary obstruction. CCA is increasing in incidence and is associated with poor prognosis.^1, 2^ Definitive therapy of CCA requires surgical resection and early diagnosis is critical for optimizing patient outcomes. Diagnostic evaluation of CCA can be accomplished via a variety of modalities; however, diagnosis of perihilar CCA and extrahepatic CCA is mostly accomplished via sampling techniques performed during endoscopic retrograde cholangiopancreatography (ERCP), such as brush cytology and transpapillary forceps biopsy.^3^

However, the early diagnosis of CCA remains an enduring challenge provided that ERCP-based sampling modalities are inaccurate for classifying biliary strictures as benign or malignant. Prior studies demonstrated that cytologic brushings and transpapillary forceps biopsies are highly specific for the classification of biliary strictures, but both techniques have poor sensitivities, ranging from 8% to 45%.^4, 5^ Despite that, the current recommendation from the American Society for Gastrointestinal Endoscopy (ASGE) is that the first line diagnostic test for biliary stricture classification is ERCP sampling via brush cytology and transpapillary forceps biopsy.^3^

Cholangioscopy is an alternative ERCP-based modality that allows for direct visualization and sampling of biliary strictures. However, the sensitivity of cholangioscopy-guided forceps biopsy is still suboptimal, approximately 70%.^6^ Prior studies have shown that physician classification of biliary strictures based solely on review of cholangioscopy footage can be accurate.^7^ However, another study demonstrated that blinded endoscopists are inaccurate and have poor interobserver variability.^8^

Thus, there remains an unmet need to develop technologies that can improve the diagnosis of biliary strictures. Emerging technologies utilizing artificial intelligence (AI) have proven capable in managing other dilemmas, such as the characterization of lesions on endoscopic ultrasound, improving the cytologic evaluation of biliary duct brushings, and the diagnosis of gastric cancer.^9-13^

Considering that it may be feasible to diagnose biliary strictures as benign or malignant using cholangioscopy footage analysis alone, it may also be possible for a cholangioscopy-based AI to address the problem of biliary stricture classification. Previously, a cholangioscopy AI was developed and validated using expert annotations of cholangioscopy footage. In that study, following retrospective review of cholangioscopy videos, the AI was found to be significantly more accurate for stricture classification (90.6%) than brush cytology (60.9%; p =0.03) or forceps biopsy (62.5%; p =0.04).^14^ These findings were further supported by a follow up, multicenter validation study which demonstrated that the cholangioscopy AI was more accurate than standard sampling techniques when exposed to recorded video data.^15^ However, both studies shared the limitation of the AI analyzing recorded cholangioscopy footage from prior ERCPs. Provided that real time application of the AI for the classification of a cholangioscopy video stream could assist in clinical adoption, it is necessary to study how the AI performs during live patient cases.^16^

Thus, the aim of this clinical trial was to assess performance of the cholangioscopy AI in real time during cholangioscopy evaluations of biliary strictures and to compare AI performance to both standard sampling techniques – brush cytology and forceps biopsy – and to blinded human evaluation of the cholangioscopy footage.

## METHODS

### Patients and Study Location

This clinical trial was approved by the UMass Chan Institutional Review Board (STUDY00000909) and was registered to clinicaltrials.gov prior to patient recruitment (NCT05779436). Development of the study design and the writing of the manuscript were performed in accordance with CONSORT-AI guidelines.^17^

This clinical trial (including patient follow up) was performed at UMass Chan Medical School from April 2023 to December 2025. Of note, the version of the AI used in this trial received no training data from UMass Chan during development.

Potential patients were approached for study inclusion in the perioperative area prior to undergoing an anticipated ERCP for evaluation of a biliary stricture that had not yet been diagnosed as benign or malignant.

For this study, patients were approached for consent if they met the following inclusion criteria: the patient age was greater than 18 years old, anticipation that the patient may undergo cholangioscopy for evaluation of a biliary stricture, and the patient could provide consent. Patients were not approached for consent if they met any of the following exclusion criteria: the patient age was less than 18 years old, the patient was a prisoner at the time of the procedure, and if the patient had a biliary stricture which had previously been established as benign or malignant.

Consented patients were included for analysis only if they underwent a cholangioscopy during the ERCP procedure and if they were found to have a biliary stricture. The decision as to whether to perform an ERCP and/or cholangioscopy was left to the discretion of the endoscopist based on their interpretation of the patient’s intraprocedural findings.

If the patient did not undergo a cholangioscopy then the patient was removed from the study, and they were not included in the final analysis. If the patient underwent an ERCP with cholangioscopy and no intrinsic biliary stricture was identified (e.g., choledocholithiasis was encountered without an accompanying stricture) the patient was not included for analysis.

Previous patients nor the public were involved in the development of this trial.

### AI Deployment

If cholangioscopy was performed during a patient’s ERCP, then the real-time cholangioscopy AI computer was positioned within view of the endoscopist. The real-time device used in this study was a custom computer that was built and coded by authors affiliated with the UMass Chan Program in Digital Medicine (PDP, MM, and NBM). The computer contained a Ryzen 7 5800x central processing unit (Advanced Micro Devices Inc, Santa Clara, California, USA) and an RTX 3900 graphics card (NVIDIA, Santa Clara, California, USA). The capture card of the real-time cholangioscopy AI computer was connected to the video output of the cholangioscopy processor (SpyGlass, Marlborough Massachusetts, USA). Once the video connections between the cholangioscopy processor and the real time computer were established, then the computer program was initiated to confirm that the cholangioscopy footage was streaming, was being processed, and was being analyzed in real time by the AI. A separate video feed of the cholangioscopy processor was also connected to the clinical video screens in the procedural room. Thus, the endoscopist had two cholangioscopy video streams to look at – one with AI processing via the real-time AI computer and one with no AI evaluation using the clinical procedure screens. A demonstration of the development and set up for the real time AI computer is provided in **Video 1**.

### ERCP and Cholangioscopy examination

Once confirmed that both the clinical and AI video screens were streaming adequately, the cholangioscope was inserted into the endoscope working channel. During this trial only the Spyglass Generation 2 cholangioscope (Boston Scientific, Marlborough Massachusetts, USA) was utilized. Endoscopists were allowed to perform cholangioscopy at their discretion. Specifically, endoscopists could perform cholangioscopy before or after alternative sampling (i.e., brush cytology or transpapillary forceps biopsy), during or after the index ERCP when a sphincterotomy had been performed, and with or without the assistance of a guidewire. The endoscopists were allowed to utilize either the clinical or AI video screens to help navigate the biliary tree. The endoscopists were not required to perform any sampling technique unless they chose to do so. If endoscopists performed cholangioscopy-guided biopsies they were not instructed that a minimum number of biopsies were required. Finally, endoscopists were allowed to sample strictures using whatever tools or techniques they felt were appropriate. Biliary tract sampling tools used during trial include a cytology brush (RX Cytology Brush, Boston Scientific, Marlborough Massachusetts, USA, transpapillary biopsy forceps (Radial Jaw 4P, Boston Scientific, Marlborough Massachusetts, USA), and cholangioscopy forceps biopsy (SpyBite Max Biopsy Forceps, Boston Scientific, Marlborough Massachusetts, USA). Only sampling procedures performed during the same ERCP where cholangioscopy AI was performed were subject to comparison and analysis.

ERCP and cholangioscopy procedures were performed by 4 endoscopists at UMass Chan Medical School. These endoscopists varied in experience (1 year to 20 years of advanced endoscopy experience) and ERCP volume (50 ERCP to 400 ERCPs performed annually).

### AI analysis

No retraining of the AI was performed prior the onset of this trial. Once cholangioscopy commenced, the real time AI computer immediately began analyzing the video stream. In real time the AI would provide inferences on a per-frame basis as to whether a cholangioscopy image appeared malignant, suspicious, benign, low-quality, or uninformative, thereby highlighting concerning areas for endoscopists to sample. Additionally, the computer would track the maximum running average AI malignancy score for the cholangioscopy procedure. This running average score was constantly re-assessed for the most recent 900 frames of cholangioscopy footage.

If the running average score crossed the pre-determined malignancy score threshold during the cholangioscopy examination that would indicate an AI prediction of malignancy (**Figure 1A-B**). If the score remained below the threshold during the examination that would indicate an AI prediction of a benign stricture. The applied score threshold was based on previously published internal validation studies. Further description of the development of the AI analysis are provided in the prior validation manuscripts.^14, 18^

**Figure 1.**
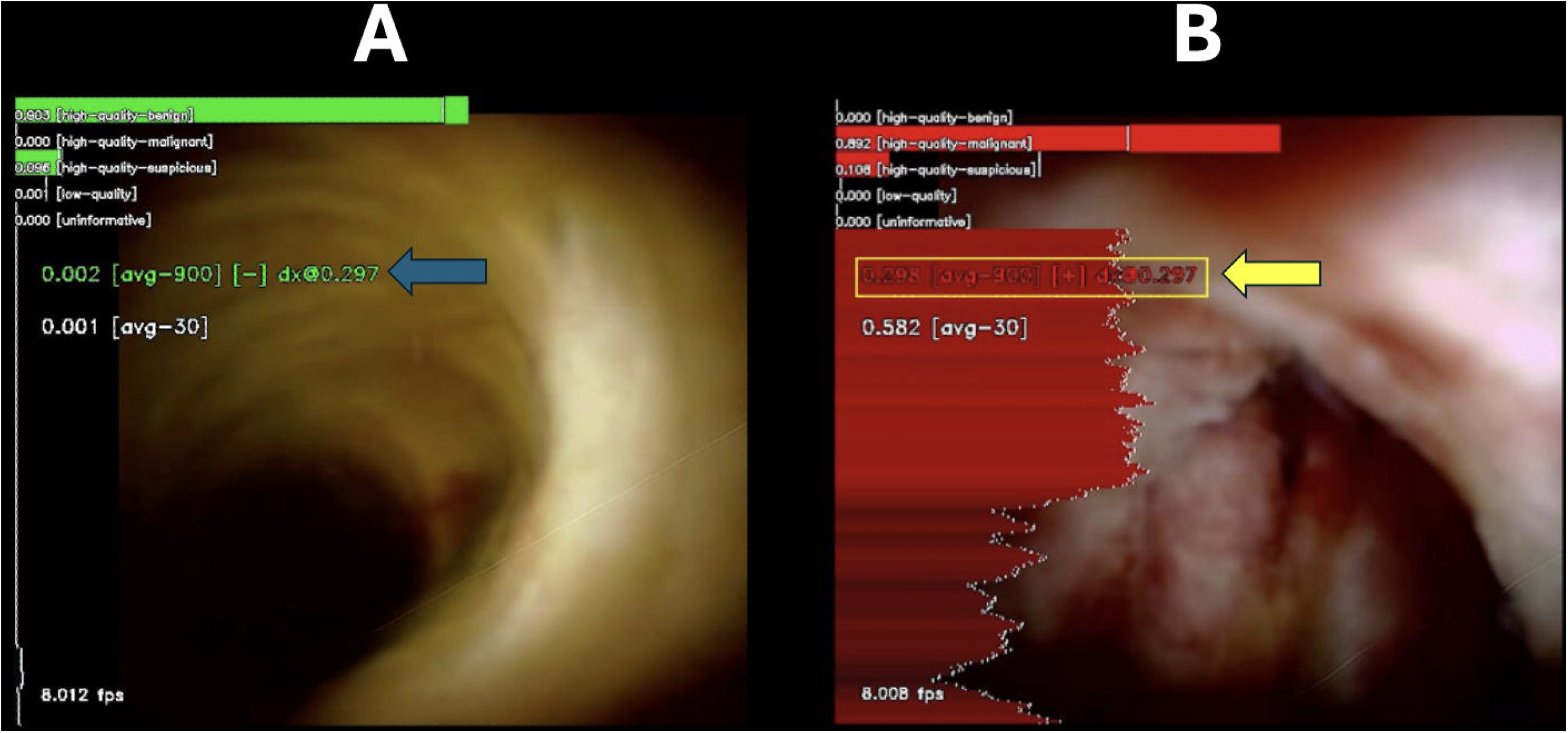
**A-B**. Panel of images demonstrating AI analysis during a patient’s cholangioscopy examination. Early during the patient’s examination, the AI-generated malignancy score was below the threshold to make a prediction for malignancy (blue arrow). Later during the patient’s examination, after the biliary stricture was adequately visualized, the malignancy score crossed the threshold (yellow arrow) indicating an AI prediction of malignancy. The AI analysis is performed in a running fashion whereby continuously the last 900 frames of data are analyzed by the AI to generate a malignancy score.

### Patient follow up and biliary stricture classification

Patients were followed for a minimum of 12 months or until a diagnosis of a malignant biliary stricture was established. We adhered to the Mayo Clinic protocol for the ultimate diagnosis of malignant biliary strictures.^19^ Specifically, a biliary stricture was classified as positive for malignancy if:

1. A malignant-appearing stricture was present on cholangiogram and…
  a. Endoscopic intraluminal brushings or tissue biopsy was positive for malignancy OR
  b. Endoscopic intraluminal brushings or tissue biopsy was strongly suspicious for cholangiocarcinoma and fluorescent in situ hybridization polysomy OR
  c. Cross-sectional imaging demonstrated the presence of a mass lesion at the site of the malignant-appearing stricture OR
  d. A CA 19-9 level of greater than 100 U/mL was present in the absence of acute bacterial cholangitis or unresolved biliary obstruction

In addition, a stricture could be considered malignant if during the 12 month follow up period cross-sectional imaging demonstrated disease progression (e.g., diagnostic imaging evidence of hepatic metastases or peritoneal carcinomatosis).

Otherwise, for a biliary stricture to be classified as benign, the stricture must not meet any of these criteria during the 12 month follow up period.

### Blinded human interpretation of cholangioscopy video recordings and video segments

At the completion of the trial, all raw cholangioscopy videos that were included for analysis were downloaded. Raw videos were edited into smaller segments containing relevant footage of biliary strictures (approximately 1-2 minutes in length). A group of advanced endoscopy fellows and advanced endoscopy attendings reviewed the footage. The fellows and attendings were provided access to the focused video segments and the entire raw footage of a procedure for assessments, if needed. The fellows and attendings were blinded to any clinical information about the patients and were only making classification predictions (i.e. negative or positive for malignancy) based on visual interpretation of videos. Fellows and attendings were not instructed to adhere to any classification criteria or guidelines when making predictions.

## STATISTICAL ANALYSIS

### Primary Outcome -- Comparison of Cholangioscopy AI to standard sampling techniques

The primary outcome of this clinical trial the diagnostic accuracy of cholangioscopy AI and standard sampling techniques – brush cytology and forceps biopsy -- for the classification of biliary strictures. Provided that the diagnostic yield of cholangioscopy-guided forceps biopsy in this trial may be influenced by the cholangioscopy AI (i.e. AI targeted sampling), separate analyses were performed – one where cholangioscopy AI was compared to the combined performance of brush cytology and transpapillary forceps biopsy (i.e. standard sampling techniques; primary outcome) and another where cholangioscopy AI was compared to the combined performance of brush cytology, transpapillary forceps biopsy, and cholangioscopy-guided forceps biopsy (secondary outcome).

When assessing combined performance of sampling techniques, if one modality was positive for malignancy, but another modality was negative, then the combined sampling would be considered positive. For example, if brush cytology was negative, but transpapillary biopsies were positive then the combination of brush cytology and biopsies would be considered positive for malignancy.

The gold standard that was used to compare cholangioscopy AI and sampling techniques was the final patient outcome after 12 months of follow up. Classification performance for cholangioscopy AI and sampling techniques included reporting sensitivity, specificity, accuracy, positive predictive value, and negative predictive value. Diagnostic accuracies of the different modalities were compared using McNemar’s test.

### Power analysis and Sample Size Calculation for the Primary Outcome

A power analysis was performed to calculate the minimum number of patients needed to detect a significant difference in the primary outcome. Previously, a retrospective validation study demonstrated that the cholangioscopy AI was 90.6% accurate for biliary stricture classification whereas brush cytology and forceps biopsy were 60.9% and 62.5% accurate, respectively. To detect a significant difference in diagnostic accuracy for biliary stricture classification, a sample size classification was performed with a power of 80% and a significance level of 0.05. Assuming a dropout rate of 15%, a minimum of 40 patients with undiagnosed biliary strictures were required for trial enrollment.

### Secondary outcomes

One secondary outcome in this trial was the comparison of cholangioscopy AI diagnostic performance to the combined diagnostic performance of brush cytology, transpapillary forceps biopsy, and cholangioscopy-guided forceps biopsy. Another secondary outcome in this trial was the comparison of AI diagnostic performance to the performance of blinded human observers. Interobserver agreement for human interpretation was calculated using Fleiss’ _K_ coefficient. Subgroup analyses were performed based on experience level. Observers who were advanced endoscopy trainees or within 1 year of graduation were considered to have junior-level experience. Observers more than 1 year out from training were considered to be “experienced.”

Like the primary outcome, assessment of all secondary outcomes included reporting sensitivity, specificity, accuracy, PPV, and NPV. Diagnostic accuracies were compared against cholangioscopy AI using McNemar’s test.

## RESULTS

### Patients

From 2023 to 2024 a total of 123 patients were approached at UMass Chan Medical School to participate in the clinical trial of which 122 provided study consent. Of the 122 patients who provided consent, 50 patients underwent cholangioscopy during their ERCP procedure. Of those patients, 9 patients were either not found to have a biliary stricture on cholangioscopy or were lost to follow up. Thus 41 patients were subsequently included in our final analysis and had chart follow up through November 2025 (**Figure 2**).

**Figure 2.**
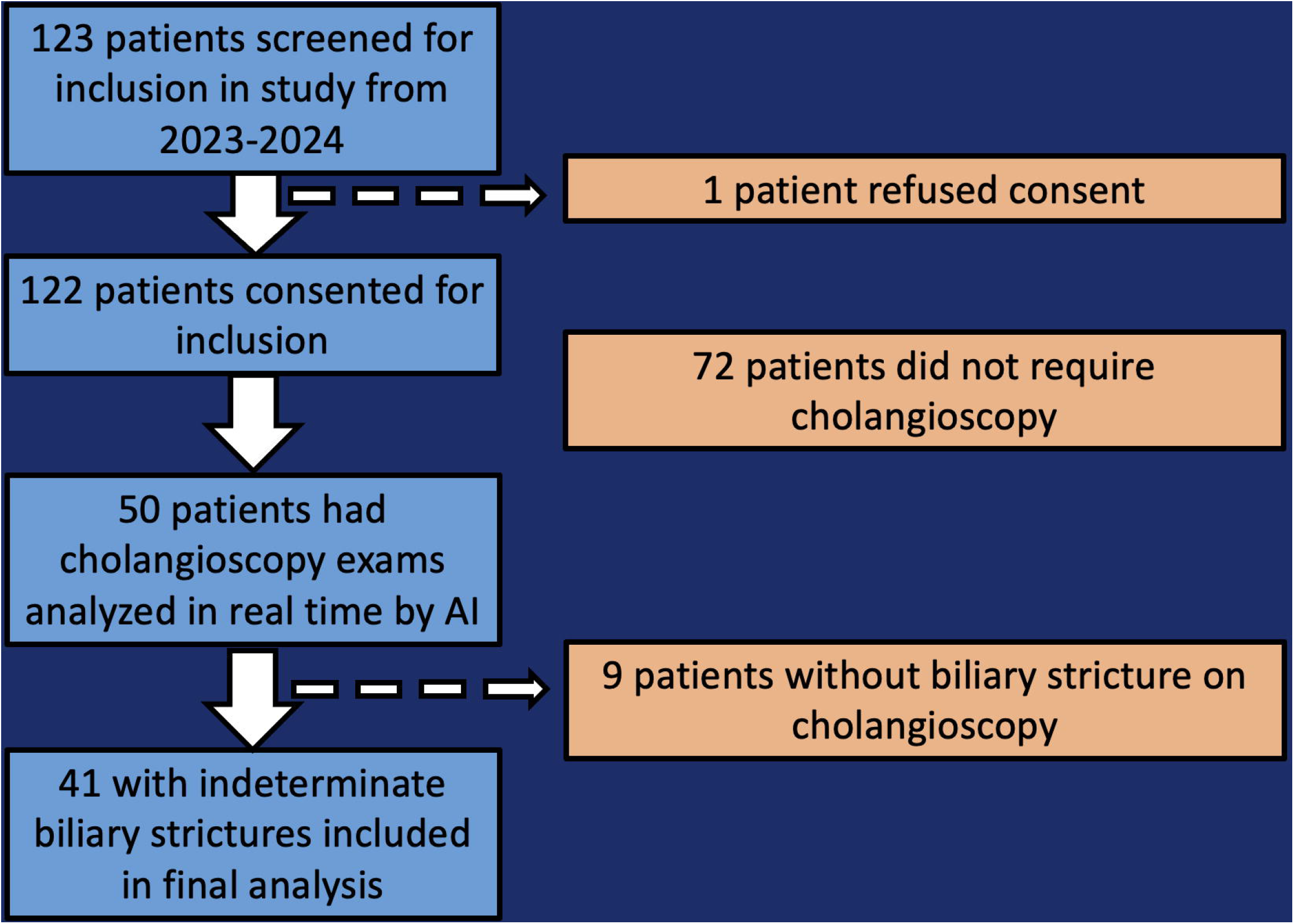
Study flow chart demonstrating the number of patients who were screened, who were consented, who underwent a cholangioscopy, and who were included in the final analysis.

Included patients were on average 65.3 ± 15.1 years old and most of the patients were male (65.9%) and non-Hispanic whites (70.7%). Most patients did not have PSC (90.2%). Several patients (43.9%) had biliary stents removed just prior to undergoing cholangioscopy.

The most common stricture location was perihilar (36.6%), followed by the distal extrahepatic duct (24.4%) and the mid extrahepatic bile duct (22.0%). In this study, 46.3% of all strictures were ultimately diagnosed as malignant. The most common diagnosis of malignant biliary strictures was cholangiocarcinoma (68.4%). Of the benign biliary strictures, most were thought to be secondary to prior choledocholithiasis (31.8%) and IgG-4 cholangiopathy (18.8%).

Additional details regarding patients are described in **Table 1**.

### Diagnostic accuracy of cholangioscopy AI and sampling techniques

AI performance analysis demonstrated that the real time system had a sensitivity of 0.895 (0.669-0.987), specificity of 0.864 (0.651-0.971), and accuracy of 0.878 (0.738-0.959) (**Figure 3**). Examples of real time AI video classification of biliary strictures are provided in **Video 2**.

**Figure 3.**
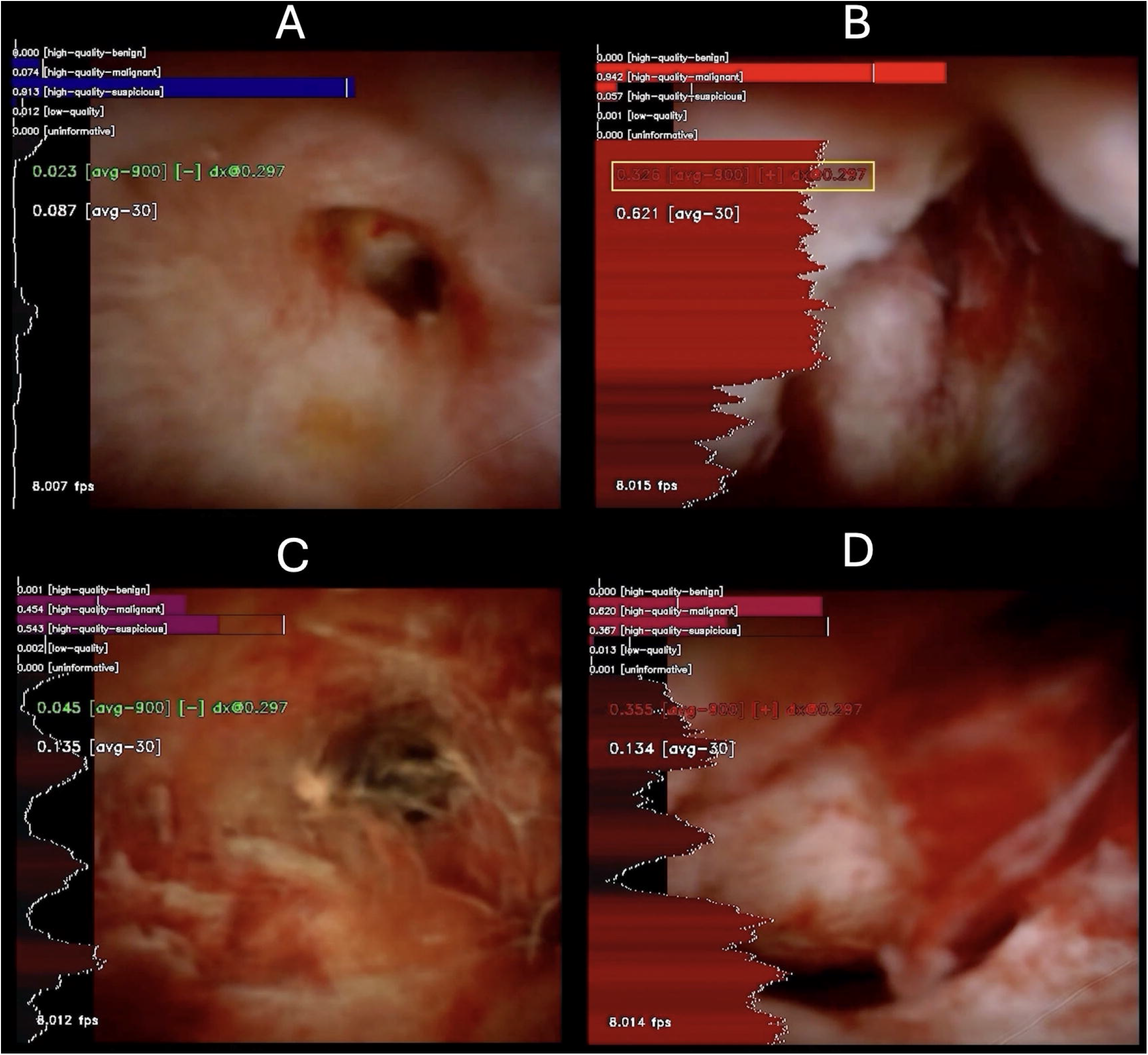
**A-D**. Panel of images demonstrating examples of both benign and malignant biliary pathologies analyzed by the AI. Panel A is a cholangioscopy image from a patient who was diagnosed with COVID-19 cholangiopathy. Panel B is a cholangioscopy image from a patient who was diagnosed with cholangiocarcinoma. Panel C is a cholangioscopy image from a patient who was diagnosed with autoimmune cholangiopathy. Panel D is a cholangioscopy image from a patient diagnosed with ovarian cancer with metastasis to the bile duct.

The combination of brush cytology and transpapillary forceps biopsy had a sensitivity of 0.438 (0.198-0.701), specificity of 1.000 (0.735-1.000), and accuracy of 0.679 (0.477-0.841). The combination of brush cytology, transpapillary forceps biopsy and cholangioscopy forceps biopsy had a sensitivity of a 0.529 (0.278-0.770), 1.000 (0.794-1.000), and accuracy of 0.758 (0.577-0.889).

For the primary outcome, the diagnostic accuracy of the AI was significantly greater than the combined performance of brush cytology and transpapillary forceps biopsy (p = 0.043).

Regarding secondary outcomes, the accuracy of the real time cholangioscopy AI was greater than the combination of all sampling techniques (brush cytology, transpapillary biopsies, and cholangioscopy biopsies), but this difference was not significant (p = 0.176)

Further details regarding performance of the AI and sampling techniques are provided in **Table 2**.

### Human performance for classification of cholangioscopy footage

A total of 14 physicians (5 junior-level, 9 experienced) from 6 community and academic centers reviewed all cholangioscopy footage from this trial.

Junior-level physicians were found to have poor interobserver agreement for classification of biliary strictures (Fleiss’ k 0.186, 95% CI 0.089-0.283), whereas experienced endoscopists had fair agreement (Fleiss’ k 0.224, 95% CI 0.173-0.275). Experienced endoscopists had greater classification than junior-level physicians, but this difference was not significant (63.1% versus 61.5%, respectively; p = 0.690). The cholangioscopy AI was significantly more accurate for biliary stricture classification than both junior-level (p = 0.001) and experienced endoscopists (p = 0.011).

Further details comparing AI performance to human observation are provided in **Table 3**.

## DISCUSSION

The classification of biliary strictures has been a difficult task for decades as sampling techniques have persistent, suboptimal accuracy. This trial demonstrates that a previously developed cholangioscopy AI can be deployed for real-time analysis and can be highly accurate for biliary stricture classification. Additionally in this trial we demonstrated that the AI is significantly more accurate in diagnosing biliary strictures than currently recommended, standard ERCP sampling techniques^3^ and is also significantly more accurate than blinded human assessment of cholangioscopy footage.

Prior studies have evaluated different cholangioscopy AIs for the diagnosis of biliary tract disease.^20-24^ This study has several key differences when compared to those prior studies. Firstly, most other cholangioscopy AIs that have been developed rely on bounding boxes for classification. The decision to avoid bounding boxes with this AI was intentional given that bounding boxes for other AI systems have raised concerns for being distracting and of limited value.^25^ Secondly, most other cholangioscopy AI studies did not limit their assessment to the evaluation of malignant and benign biliary strictures, but also included benign cases where no strictures were present, which could result in an overestimation of AI performance. Provided that the key dilemma is the differentiation of malignant biliary strictures from benign strictures, this trial excluded any cholangioscopy case where an intrinsic biliary stricture was not identified. Thirdly, like many prior endoscopy-based AIs, most cholangioscopy AIs have only been tested on retrospective datasets and have not been validated for real-time clinical deployment. In this trial, we developed and deployed cholangioscopy AI-based technology for real-time assessment of biliary strictures during ERCP with a 12 month-follow up period. The diagnostic accuracy of the real time cholangioscopy AI during this trial (88.2%) closely resembled the accuracy of the AI when it had been tested previously on retrospective video datasets from two validation studies (90.6% and 85.3%, respectively). Additionally, this trial entirely took place at a center that did not provide any training data during the AI’s development. Importantly, this suggests that the cholangioscopy AI performance is consistent and reliable, even when deployed in real-time.

In this trial, the cholangioscopy AI significantly outperformed brush cytology and transpapillary forceps biopsy for the task of stricture classification. The AI had greater accuracy than the combination of brush cytology, transpapillary forceps biopsy, and cholangioscopy forceps biopsy; however, this difference was not statistically significant. The accuracy of cholangioscopy forceps biopsy in this study was greater than what was seen in both prior validation studies as well as the retrospective cohort that was used for comparison in this trial. The real-time AI system does provide guidance as to where endoscopists should sample within the bile duct. Thus, this improvement in yield may suggest that AI-assisted cholangioscopy forceps biopsy improves sampling; however, this trial was underpowered to make that comparison and requires additional study.

In addition to outperforming standard, first-line sampling techniques, the real-time AI significantly outperformed both junior-level and experienced endoscopists for stricture classification. Additionally, this trial demonstrated that both fellows and experienced endoscopists only had poor or fair interobserver agreement and experienced endoscopists did not have significantly superior performance when compared to fellows. These findings have important implications. Firstly, our findings are consistent with that of recent analyses which suggest that blinded physicians have suboptimal interobserver agreement and accuracy when assessing cholangioscopy videos of strictures.^8^ Secondly, human cholangioscopy experience is not clearly associated with improved performance in stricture classification. Thirdly, the AI may have a role in assisting endoscopists in detecting malignancy, regardless of their experience level.

While our trial has strengths it also has limitations. Firstly, while this trial occurred at a center that did not provide any training data for AI development, this trial was only performed at a single center. Thus, it is not clear if the AI will have consistent performance in various study populations where different stricture phenotypes are more common, such as primary sclerosing cholangitis. Secondly, in this trial we allowed endoscopists to perform biliary stricture sampling at their discretion and did not recommend a standard sampling process (e.g., number of brush passes to perform, number of biopsies to perform with forceps). Thus, performance of sampling modalities may be heterogeneous and partially dependent on the practice of the performing provider. However, by allowing endoscopists to perform sampling techniques at their discretion, this study captures real world assessment practices of biliary strictures. Thirdly, the AI developed in this trial has only ever been validated on a single type of cholangioscope. It is unclear if the AI can be accurate for stricture classification when used with other cholangioscopes.

This trial reports on the assessment of real time deployment of a cholangioscopy AI for biliary stricture classification. While the AI was more accurate than standard sampling techniques and human assessment, it remains unclear what the future role cholangioscopy AI in the management of biliary strictures is. It is unlikely that AI will completely replace other diagnostic tests but may instead be part of an ensemble of a variety of diagnostic techniques for biliary strictures. In the future, multicenter trials are needed to reinforce the findings from this trial and to elucidate how cholangioscopy AI could be used to improve the care of patients with biliary strictures.

## Data Availability

Data Availability Statement: All data produced in the present study are available upon reasonable request to the authors.

## Acronyms

CCA: Cholangiocarcinoma
ERCP: Endoscopic retrograde cholangiopancreatography
ASGE: American Society of Gastrointestinal Endoscopy
AI: Artificial intelligence

---

**Video 1**. Video providing details regarding the development and set up of the real time cholangioscopy AI computer used during the clinical trial.

**Video 2**. Video providing examples of real time AI deployment for four cases (two benign, two malignant) during the clinical trial.

